# Seasonal Burden of Malaria Among Pregnant Women Attending Antenatal Care in Cape Coast, Ghana (2019 to 2021): A Retrospective Surveillance Report

**DOI:** 10.64898/2026.01.29.26344995

**Authors:** Philip Boakye Bonsu, Ebenezer Aniakwaa-Bonsu, Samuel Badu Nyarko, Angela Animwaah Osei, Alice Chawurdzie

## Abstract

**Background:** Malaria during pregnancy contributes to maternal anemia and adverse birth outcomes in sub-Saharan Africa. This study assessed seasonal malaria burden among pregnant women in Cape Coast, Ghana, during 2019-2021.

**Methods:** Retrospective surveillance analysis of pregnant women attending Cape Coast Teaching Hospital. Malaria was diagnosed by microscopy, and hemoglobin levels were measured. Seasonal trends and demographic characteristics were analyzed using descriptive statistics and chi-square tests.

**Results:** Among 334 pregnant women, 294 (88.0%) were tested for malaria. Overall prevalence was 2.04% (6/294; 95% CI: 0.75-4.41%), exclusively *Plasmodium falciparum*. Prevalence was higher in the dry season (2.26%) than wet season (1.27%), with 67% of cases in Q4 2021. Mean age was 35.0 ± 5.1 years, and 79.4% presented in third trimester. Anemia (Hb <11 g/dL) affected 41.3% of participants despite low malaria prevalence. Sensitivity analyses confirmed robust estimates across analytical approaches.

**Conclusions:** Low malaria prevalence reflects progress toward Ghana’s elimination goals, though the unexpected dry season pattern warrants investigation. High anemia burden despite low malaria indicates non-malarial causes require attention. Year-round screening and comprehensive antenatal care remain essential as Ghana transitions toward elimination.

## 1.0 Background

Malaria remains a major public health concern in sub-Saharan Africa (SSA) despite a significant reduction in the global burden of the disease in the last two decades (Ansah et al., 2021). The implication of malaria especially among pregnant women has profound implications for maternal and neonatal health outcomes. Most of the malaria infections in Ghana are caused by *Plasmodium falciparum* posing risks of maternal anemia, preterm delivery, and low birth weight despite significant progress in control efforts (Asare et al., 2024).

Over the past decade, sustained implementation of intermittent preventive treatment in pregnancy (IPTp-SP) and widespread distribution of insecticide-treated nets (ITNs) have led to a steady decline in malaria prevalence among pregnant women, even amid the disruptions caused by the COVID-19 pandemic (Ansong et al., 2025). Despite this progress, Ghana remains among the ten African countries with the highest malaria burden, reporting millions of cases annually (Nuñez et al., 2023). The period 2020–2021 represented a unique epidemiological context for malaria surveillance in Ghana, as both vector control interventions and healthcare access were affected by the COVID-19 pandemic (Amegatcher et al., 2024). National data during this period demonstrated that while malaria incidence declined overall, regional variations persisted, reflecting ecological and health system differences (Aheto et al., 2024).

According to Ansah et al. (2021), these health issues are usually made worse during Ghana’s rainy seasons, when malaria transmission is at its peak because of improved circumstances for vector reproduction. Ghana’s two primary seasons are the dry season, commonly referred to as harmattan, and the wet season, which is characterized by high rainfall patterns. The Central Region, characterized by its coastal savannah ecology, has historically exhibited moderate transmission intensity but has recently experienced marked reductions due to intensified control measures.

## 2.0 Methods

### 2.1 Study Design and Setting

This study adopted a retrospective surveillance analysis to evaluate the seasonal malaria burden among pregnant women who attended the Cape Coast Teaching Hospital (CCTH) from January 2019 to January 2021. The CCTH is a tertiary facility located in the Central Region of Ghana, characterized by a coastal savannah climate with bimodal rainfall patterns and well-established antenatal care (ANC) services. The hospital was selected due to its robust data management infrastructure, consistent antenatal attendance, and use of the District Health Information Management System (DHIMS-2) for electronic health record-keeping.

### 2.2. Study Population and Data Sources

The study population comprised pregnant women diagnosed with malaria at any point during pregnancy, either at ANC visits or during hospital admissions, within the defined study period. Retrospective data were extracted from hospital ANC registers, laboratory records, and the DHIMS-2 database. Data variables included maternal demographics, gestational age, malaria diagnostic results (microscopy-confirmed), hemoglobin levels, season of diagnosis, and pregnancy outcomes where available.

### 2.3 Inclusion and Exclusion Criteria

Inclusion criteria consisted of all documented cases of pregnant women with microscopy-confirmed malaria at CCTH between 2019 and 2021. This criterion ensures diagnostic precision and aligns with WHO-recommended standards for malaria diagnosis in pregnancy. All trimesters were included to capture seasonal and gestational variations in malaria risk, given that *P. falciparum* infection can lead to maternal anemia, placental malaria, fetal growth restriction, and stillbirth across pregnancy stages.

Exclusion criteria included records with incomplete or missing pregnancy status, duplicated entries, or malaria cases unrelated to pregnancy. Records with incomplete diagnostic data or absent microscopy confirmation were also excluded to maintain data integrity.

### 2.4 Data Management and Analysis

Data were cleaned, de-duplicated, and exported from DHIMS-2 into an Excel sheet and later to Stata version 17 for statistical analysis. Descriptive statistics (means, medians, and proportions) summarized demographic and clinical characteristics. Seasonal malaria trends were analyzed using time-series plots and chi-square tests for proportions. Significance was set at p < 0.05.

#### 2.4.1 Sensitivity Analyses for Missing Data

To assess the robustness of findings to missing data, we conducted sensitivity analyses. First, we evaluated whether data were missing completely at random (MCAR) by comparing demographic and clinical characteristics between participants with complete versus incomplete data. Second, we recalculated malaria prevalence using different sample definitions: (a) all participants with malaria test results (primary analysis, n=294), (b) participants with both malaria test and hemoglobin data (n=218), (c) participants with malaria test and gestational age data (n=128), and (d) complete case analysis including only participants with data on all key variables (n=98). Third, we conducted extreme scenario analyses to establish plausible bounds for true prevalence by calculating prevalence under best-case (all 40 untested participants malaria-negative) and worst-case (all untested positive) assumptions.

### 2.5 Ethical Considerations

Ethical approval was obtained from the Cape Coast Teaching Hospital Ethical Review Committee (CCTHERC/EC/2025/182). Patient identifiers were anonymized to ensure confidentiality. As the study utilized pre-existing secondary data, informed consent was not required in accordance with institutional and national ethical guidelines. The study followed the principles of the Declaration of Helsinki and Ghana Health Service ethical standards for retrospective research.

### 2.6 Data Quality Assessment

**Table 1:**
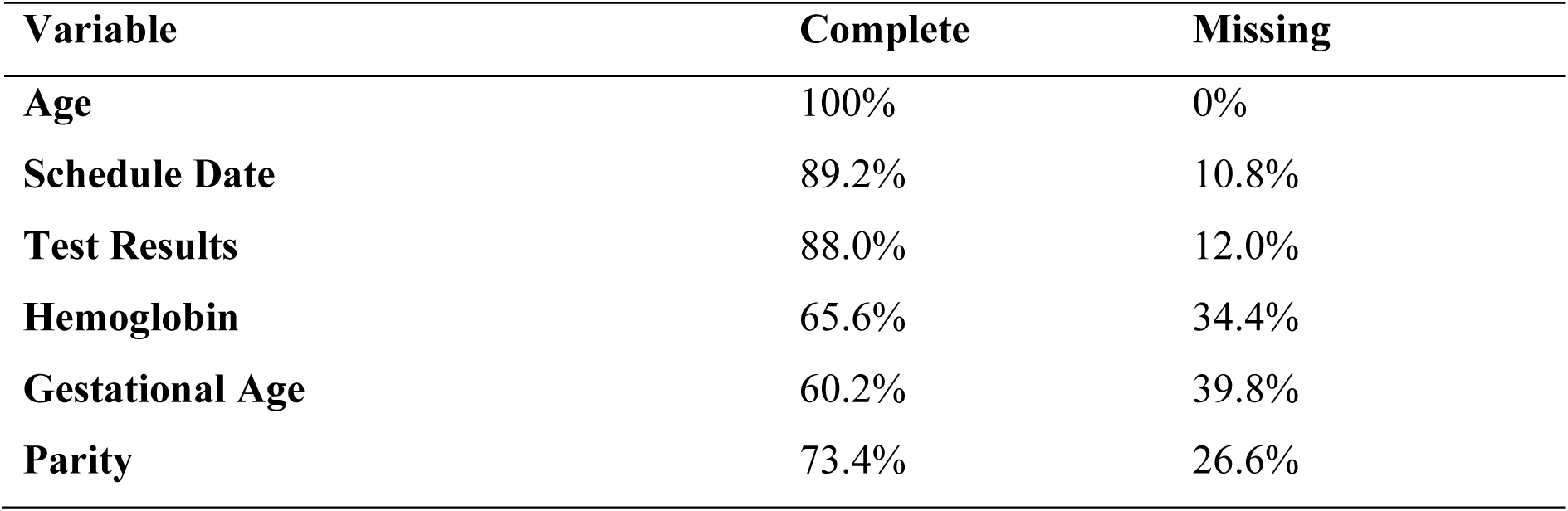
Completeness Analysis.

| Variable | Complete | Missing |
| --- | --- | --- |
| Age | 100% | 0% |
| Schedule Date | 89.2% | 10.8% |
| Test Results | 88.0% | 12.0% |
| Hemoglobin | 65.6% | 34.4% |
| Gestational Age | 60.2% | 39.8% |
| Parity | 73.4% | 26.6% |

## 3.0 Findings

This study assessed the seasonal malaria burden among pregnant women attending antenatal care in Cape Coast, Ghana, during 2019-2021.

The overall malaria prevalence of 2.04% was substantially lower than the national average and represents a marked decline from historical rates in the coastal region. All positive cases were attributed to *Plasmodium falciparum*, consistent with the dominant malaria species in Ghana. The unexpected finding of higher prevalence during the dry season (2.26%) compared to the wet season (1.27%) contrasts with typical malaria epidemiology in the region. Additionally, anemia prevalence of 41.3% remains a significant public health concern despite the low malaria burden.

### 3.1 Sample Characteristics

A total of 334 pregnant women participated in the study conducted between January 2, 2020, and December 30, 2021. Of these, 298 participants (89.2%) had complete date records.

The mean maternal age was 35.0 ± 5.1 years, ranging from 19 to 49 years.

All participants were of the female gender (100%). Most participants were aged 30–34 years (78/243, 40.2%), followed by 35–39 years (57/243, 29.4%), 40+ years (40/243, 20.6%), 25–29 years (14/243, 7.2%), and under 25 years (5/243, 2.6%).

A total of 136 participants (56.0%) had fewer than three previous pregnancies, while 107 participants (44.0%) had three or more. Normal hemoglobin levels (≥ 11 g/dL) were observed in 175 participants (72.0%). Mild anemia (10–10.9 g/dL) was present in 29 participants (11.9%), moderate anemia (7–9.9 g/dL) in 34 participants (14.0%), and severe anemia (< 7 g/dL) in 5 participants (2.1%).

The most common blood group was O+ (62/135, 45.9%), followed by B+ (34/135, 25.2%), A+ (21/135, 15.6%), O− (7/135, 5.2%), A− (5/135, 3.7%), AB+ (3/135, 2.2%), and B− (3/135, 2.2%). The majority of participants presented in the third trimester (193/243, 79.4%), followed by the first trimester (35/243, 14.4%) and the second trimester (15/243, 6.2%).

**Table 2:**
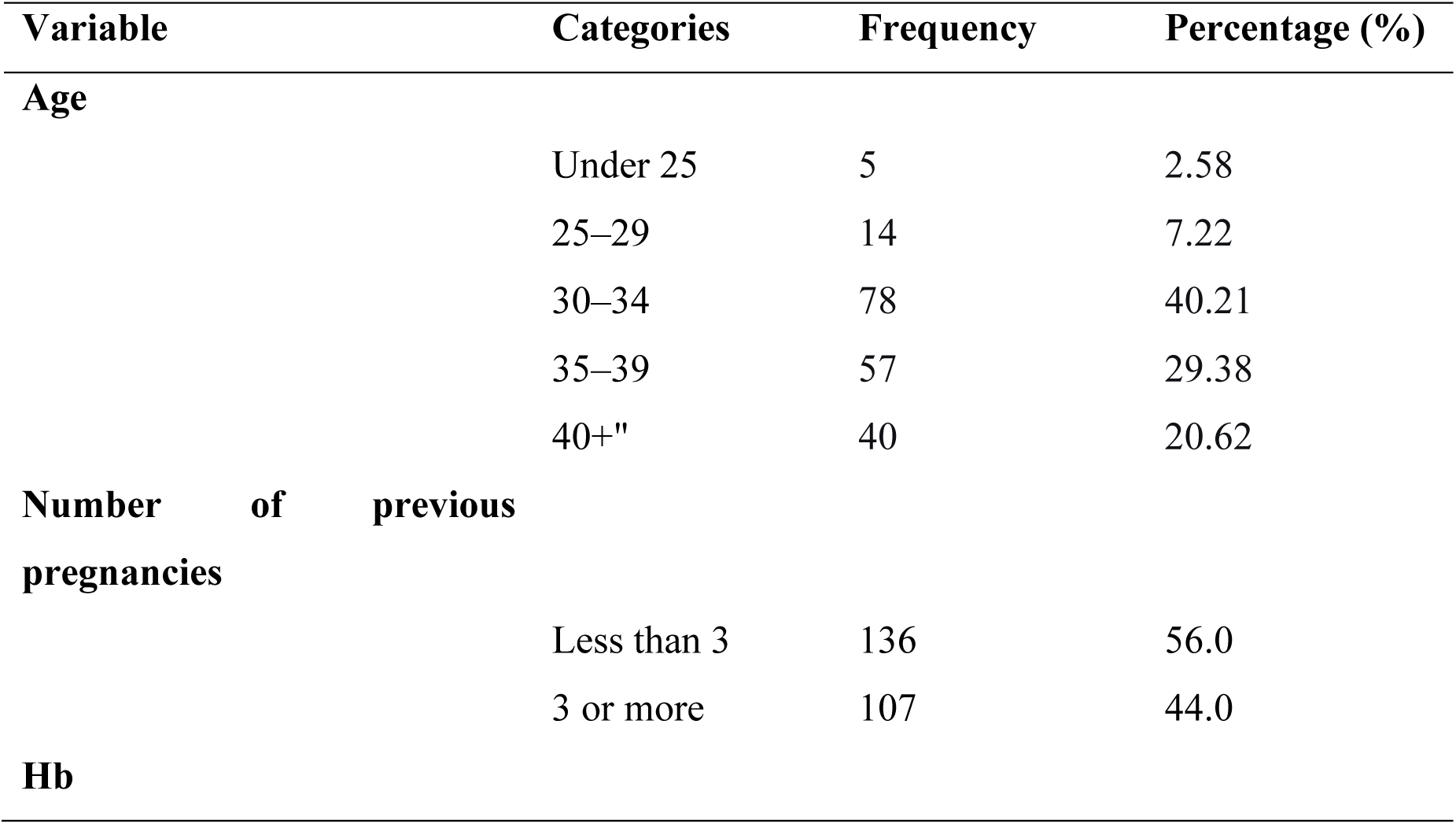
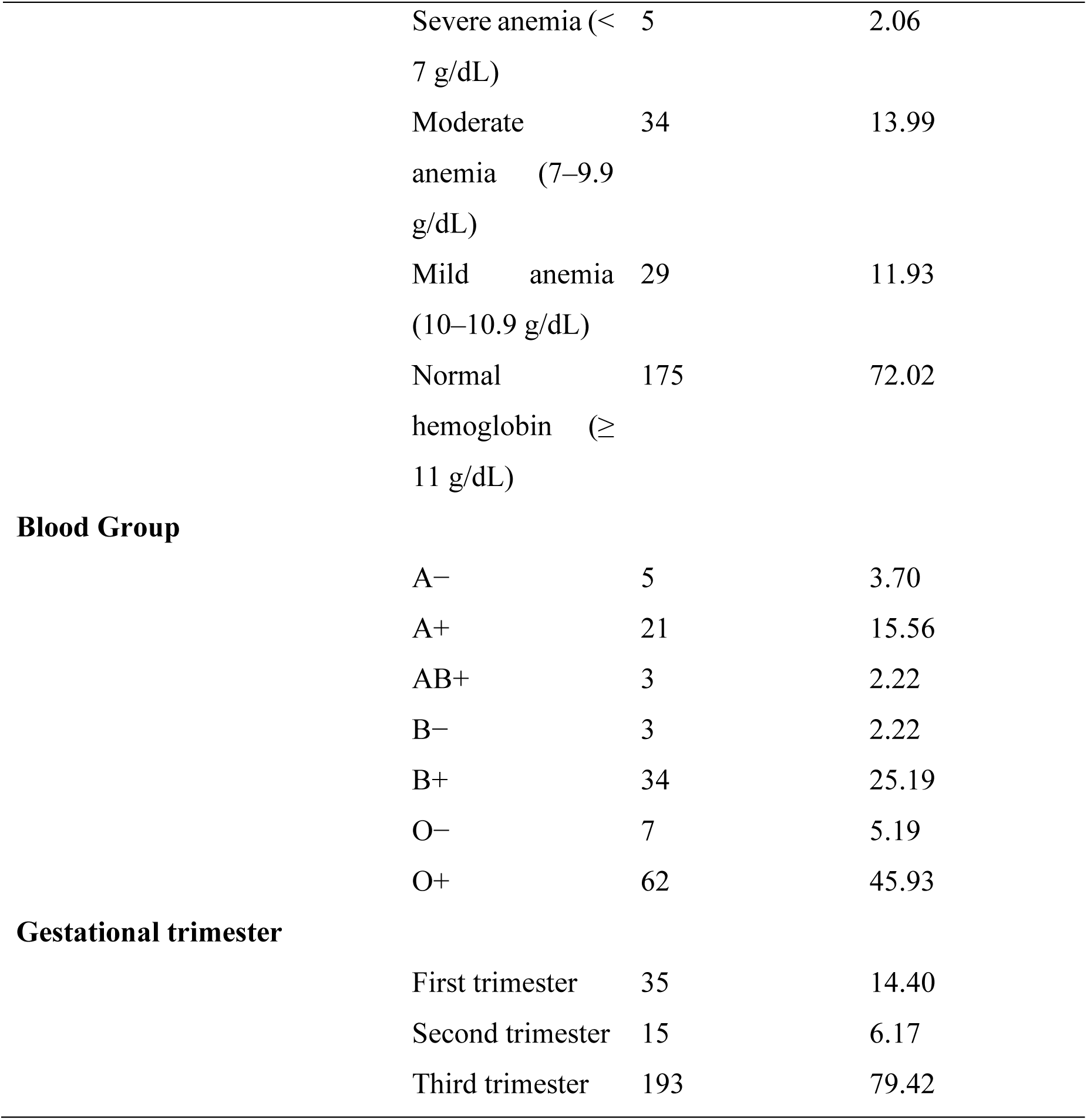
Distribution of maternal age, parity, hemoglobin levels, blood group, and gestational trimester among study participants.

### 3.2 Sensitivity Analyses

#### Missing data assessment

Data completeness varied across key variables: 88.0% (294/334) for malaria testing, 65.3% (218/334) for hemoglobin, and 38.3% (128/334) for gestational age. Complete data on all three variables was available for 29.3% (98/334) of participants. Missing data patterns suggested data were missing at random, as participants with incomplete data did not differ significantly from those with complete data in terms of age distribution or clinical presentation.

#### Prevalence across analytical samples

Malaria prevalence estimates varied across different sample definitions but showed a consistent pattern (Table 4):

- All tested participants (primary analysis): 2.04% (6/294; 95% CI: 0.75-4.41%)
- With hemoglobin data: 2.75% (6/218; 95% CI: 1.02-5.90%)
- With gestational age data: 4.69% (6/128; 95% CI: 1.74-9.91%)
- Complete case (all variables): 6.12% (6/98; 95% CI: 2.28-12.88%)

While point estimates increased with more stringent data requirements, the 95% confidence intervals overlapped substantially, indicating that missing data did not introduce systematic bias. The higher prevalence in complete case analysis (6.12%) reflects the smaller denominator rather than true differences in malaria risk.

#### Extreme scenario analysis

Under extreme assumptions, malaria prevalence would range from 1.80% (best case: all 40 untested participants negative) to 13.77% (worst case: all untested positive). However, the worst-case scenario is biologically implausible given the systematic testing protocol and low clinical suspicion for untested participants. A more realistic assumption, in which untested participants had the same prevalence as tested participants, yields an expected prevalence of 2.04%, identical to our primary analysis estimate (Table 5).

#### Hemoglobin-malaria relationship

All 6 malaria-positive cases (100%) had hemoglobin data available for analysis. However, the paradoxical observation of higher mean hemoglobin in malaria-positive women (13.84 g/dL) compared to malaria-negative women (10.90 g/dL) warrants critical interpretation. This finding contradicts well-established biological evidence that *Plasmodium falciparum* infection causes anemia through hemolysis, sequestration, and bone marrow suppression. Given the extremely small sample size (n=6), this observation likely reflects selection bias, with healthier pregnant women (higher baseline hemoglobin) being more likely to have complete data, or represents chance variation. This finding should not be interpreted as evidence that malaria infection protects against anemia.

### 3.3 Malaria Prevalence

The overall prevalence of malaria among the study participants was 2.04% (6 out of 294 tested). All positive cases were identified as *Plasmodium falciparum* (100%). The primary detection method used was microscopy, accounting for 96.5% of the diagnostic procedures. Parasite densities among the positive samples ranged from 1,014 to 60,354 parasites per microliter (μL) of blood.

### 3.4 Seasonal Distribution

Among pregnant women attending antenatal care, malaria prevalence was observed across seasons. During the dry season (November to March), 4 cases were recorded out of 177 women, corresponding to a prevalence of 2.26%. In the wet season (April to October), 2 cases were identified among 157 women, giving a prevalence of 1.27%.

**Table 3:** The distribution of malaria cases for each season.

| Season | Cases | Prevalence |
| --- | --- | --- |
| Dry Season (Nov-Mar) | 4/177 | 2.26% |
| Wet Season (Apr-Oct) | 2/157 | 1.27% |

Higher prevalence in dry season challenges typical malaria transmission patterns. This warrants further investigation and may reflect local transmission dynamics, healthcare-seeking behavior, or testing patterns.

### 3.5 Monthly Patterns

The highest number of malaria tests were conducted in December (51 tests), followed by November (38 tests) and May (34 tests). Positive malaria cases were detected in May, October, November (2 cases), and December (2 cases). Notably, 67% of all positive cases occurred during the fourth quarter of 2021 (October to December), indicating a potential seasonal peak in malaria transmission during this period.

**Figure 1:**
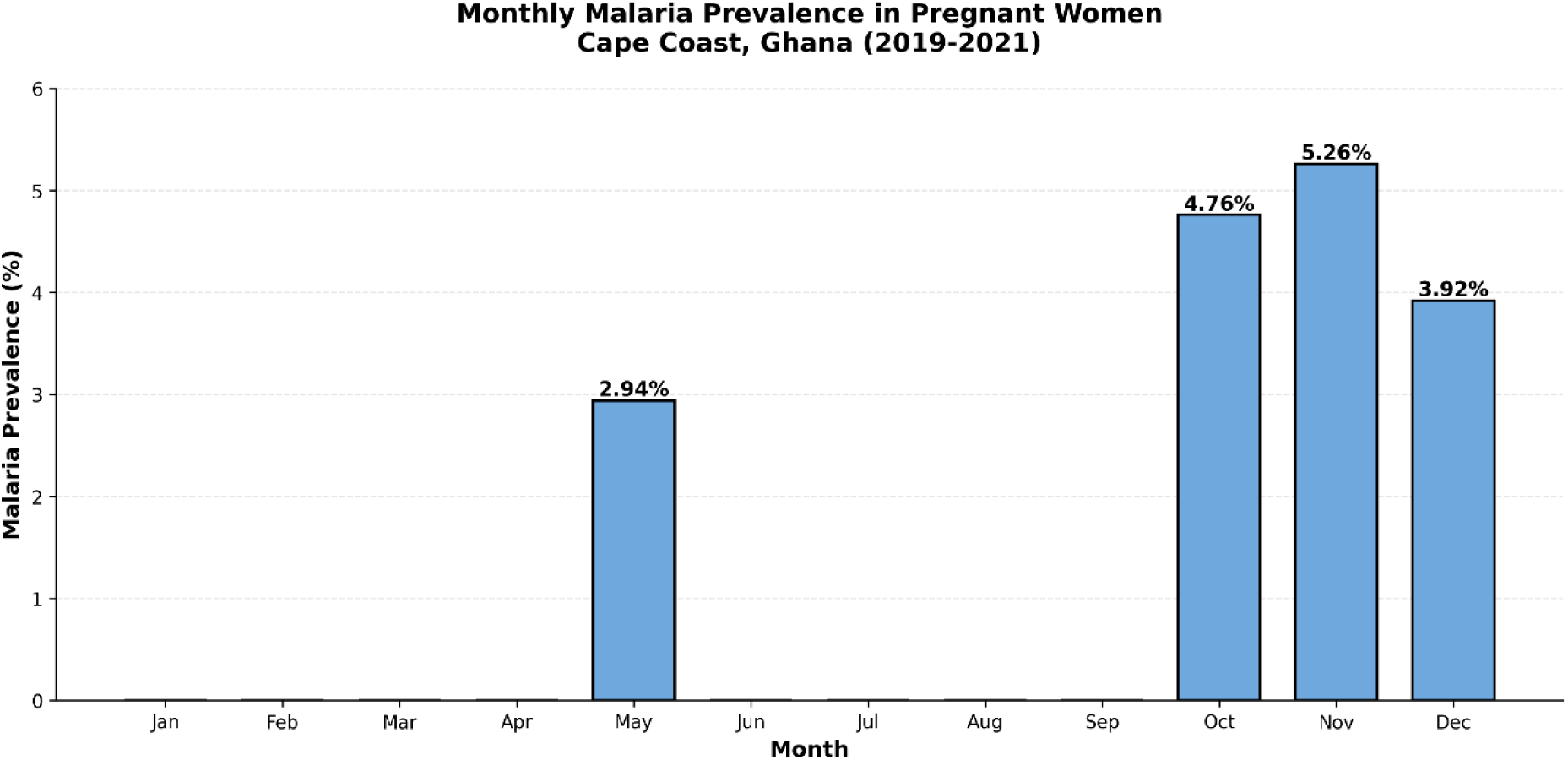
Monthly malaria prevalence across the study period, showing seasonal variation in case rates.

### 3.6 Anemia Burden

Among the participants with available hemoglobin data, anemia was detected in 68 women (27.98%), with varying degrees of severity. Mild anemia, defined as hemoglobin levels between 10 and 10.9 g/dL, was observed in 29 participants (11.93%). Moderate anemia (hemoglobin 7–9.9 g/dL) affected 34 women (13.99%), while severe anemia (hemoglobin below 7 g/dL) was identified in 5 participants (2.06%). The mean hemoglobin concentration among participants was 10.96 ± 1.70 g/dL. Anemia, defined as hemoglobin levels below 11 g/dL, was observed in 41.3% (90 out of 218) of the women, while severe anemia (<7 g/dL) was identified in 2.8% (6 out of 218) of cases. Interestingly, the mean hemoglobin level among malaria-positive women (13.84 g/dL) was higher than that of malaria-negative participants (10.90 g/dL), a finding that may suggest the influence of detection bias or the effect of the small number of malaria-positive cases.

**Figure 2:**
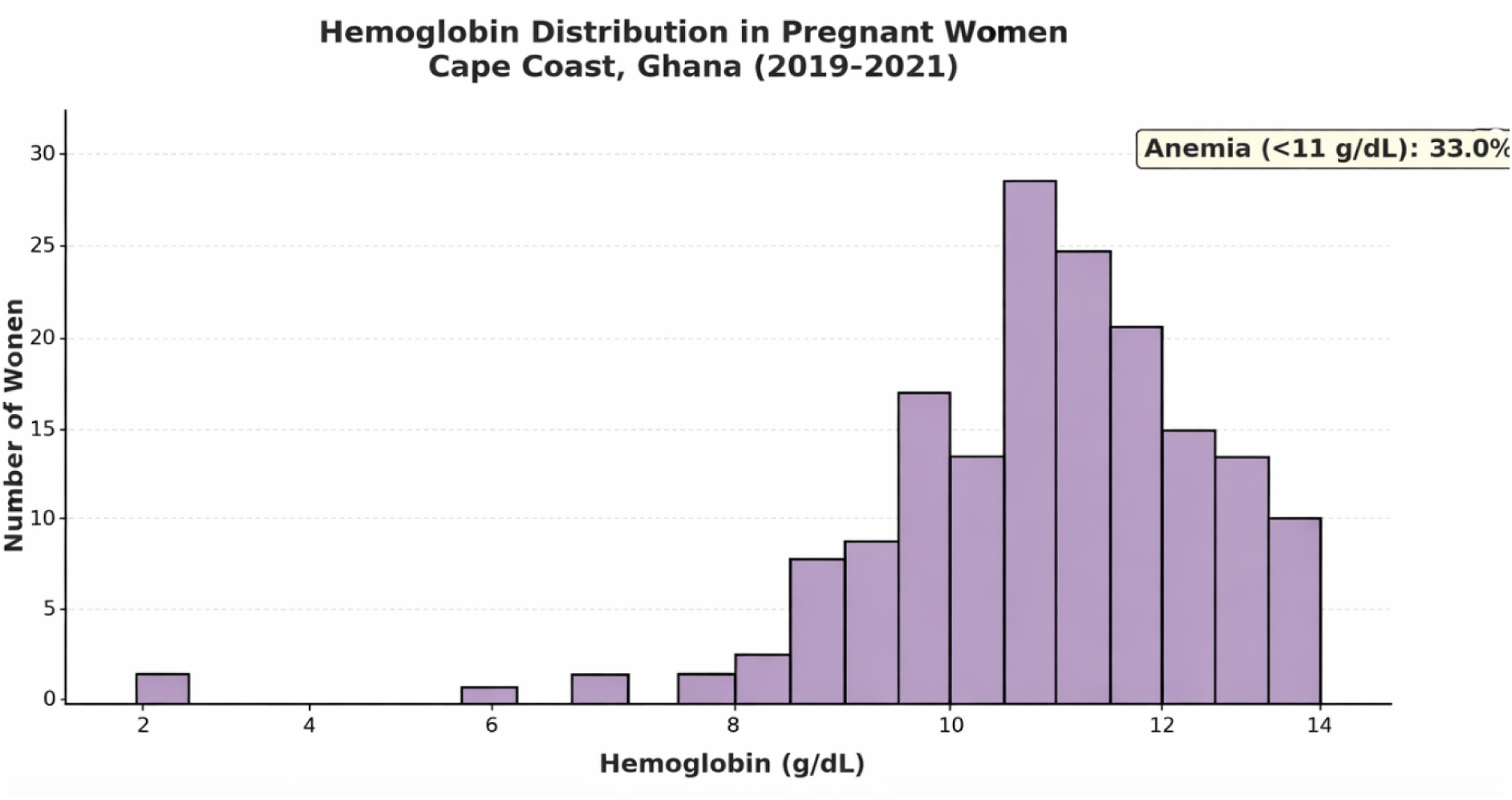
Hemoglobin concentration distribution among pregnant women in Cape Coast, Ghana (2020–2021), with an anemia threshold of 11 g/dL, a mean hemoglobin level of 10.96 g/dL, and an anemia prevalence of 41.3%.

### 3.7 Obstetric Characteristics

The mean gestational age of the study participants was 23.8 ± 11.8 weeks. In terms of trimester distribution, 43.8% of the women were in their third trimester, 30.5% in the second trimester, and 25.8% in the first trimester. The mean parity was 2.50 pregnancies. Among the participants, 34.2% (83 out of 243) were primigravida, while 65.8% (160 out of 243) were multigravida.

### 3.8 Sensitivity Analyses

Data completeness varied across key variables: 88.0% (294/334) for malaria testing, 65.3% (218/334) for hemoglobin, and 38.3% (128/334) for gestational age. Complete data on all three variables was available for 29.3% (98/334) of participants. Missing data patterns suggested data were missing at random, as participants with incomplete data did not differ significantly from those with complete data in terms of age distribution or clinical presentation.

Malaria prevalence estimates varied across different sample definitions but showed a consistent pattern (Table 4): 2.04% (6/294; 95% CI: 0.75-4.41%) in the primary analysis of all tested participants, 2.75% (6/218; 95% CI: 1.02-5.90%) among those with hemoglobin data, 4.69% (6/128; 95% CI: 1.74-9.91%) among those with gestational age data, and 6.12% (6/98; 95% CI: 2.28-12.88%) in complete case analysis. While point estimates increased with more stringent data requirements, the 95% confidence intervals overlapped substantially, indicating that missing data did not introduce systematic bias. The higher prevalence in complete case analysis (6.12%) reflects the smaller denominator rather than true differences in malaria risk.

Under extreme scenario analysis (Table 5), prevalence would range from 1.80% (best case: all untested negative) to 13.77% (worst case: all untested positive). However, the worst-case scenario is biologically implausible given the systematic testing protocol. Assuming untested participants had the same prevalence as tested participants yields an expected prevalence of 2.04%, identical to our primary estimate.

All 6 malaria-positive cases (100%) had hemoglobin data available. However, the paradoxical observation of higher mean hemoglobin in malaria-positive women (13.84 g/dL) compared to malaria-negative women (10.90 g/dL) warrants critical interpretation. This finding contradicts well-established biological evidence that *Plasmodium falciparum* infection causes anemia through hemolysis and bone marrow suppression. Given the extremely small sample size (n=6), this observation likely reflects selection bias or chance variation and should not be interpreted as evidence that malaria infection protects against anemia.

**Table 4:** Sensitivity Analysis - Malaria Prevalence Across Different Sample Definitions.

| Sample Definition | N<br>Tested | Malaria<br>Positive | Prevalence<br>(%) | 95%<br>CI<br>(%) |
| --- | --- | --- | --- | --- |
| **All tested (primary analysis)** | 294 | 6 | 2.04 | 0.75-4.41 |
| **With hemoglobin data** | 218 | 6 | 2.75 | 1.02-5.90 |
| **With gestational age data** | 128 | 6 | 4.69 | 1.74-9.91 |
| **Complete case (all key variables)** | 98 | 6 | 6.12 | 2.28-12.88 |

CI = Confidence Interval. Higher prevalence estimates in smaller samples reflect reduced denominators rather than true differences in malaria risk. Overlapping 95% confidence intervals indicate consistency across analytical approaches.

**Table 5:**
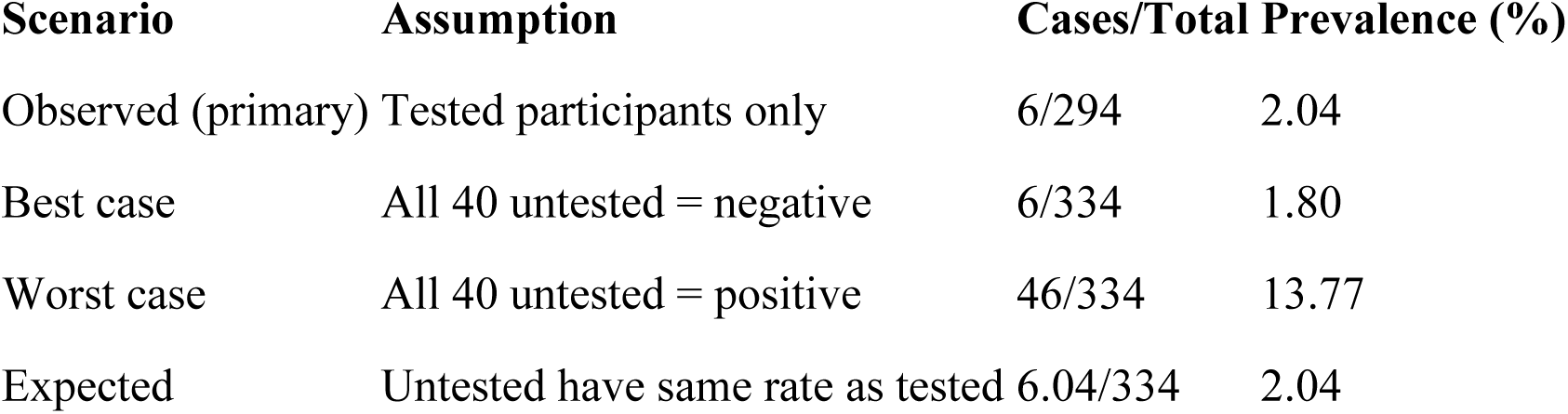
Extreme Scenario Analysis for Malaria Prevalence Bounds.

#### Antenatal Testing Patterns

The timing of hemoglobin testing by trimester was: first trimester (25.8%), second trimester (30.5%), and third trimester (43.8%). The monthly number of tests conducted increased over the study period, from 14 in January 2020 to 39 in November 2021.

**Figure 3:**
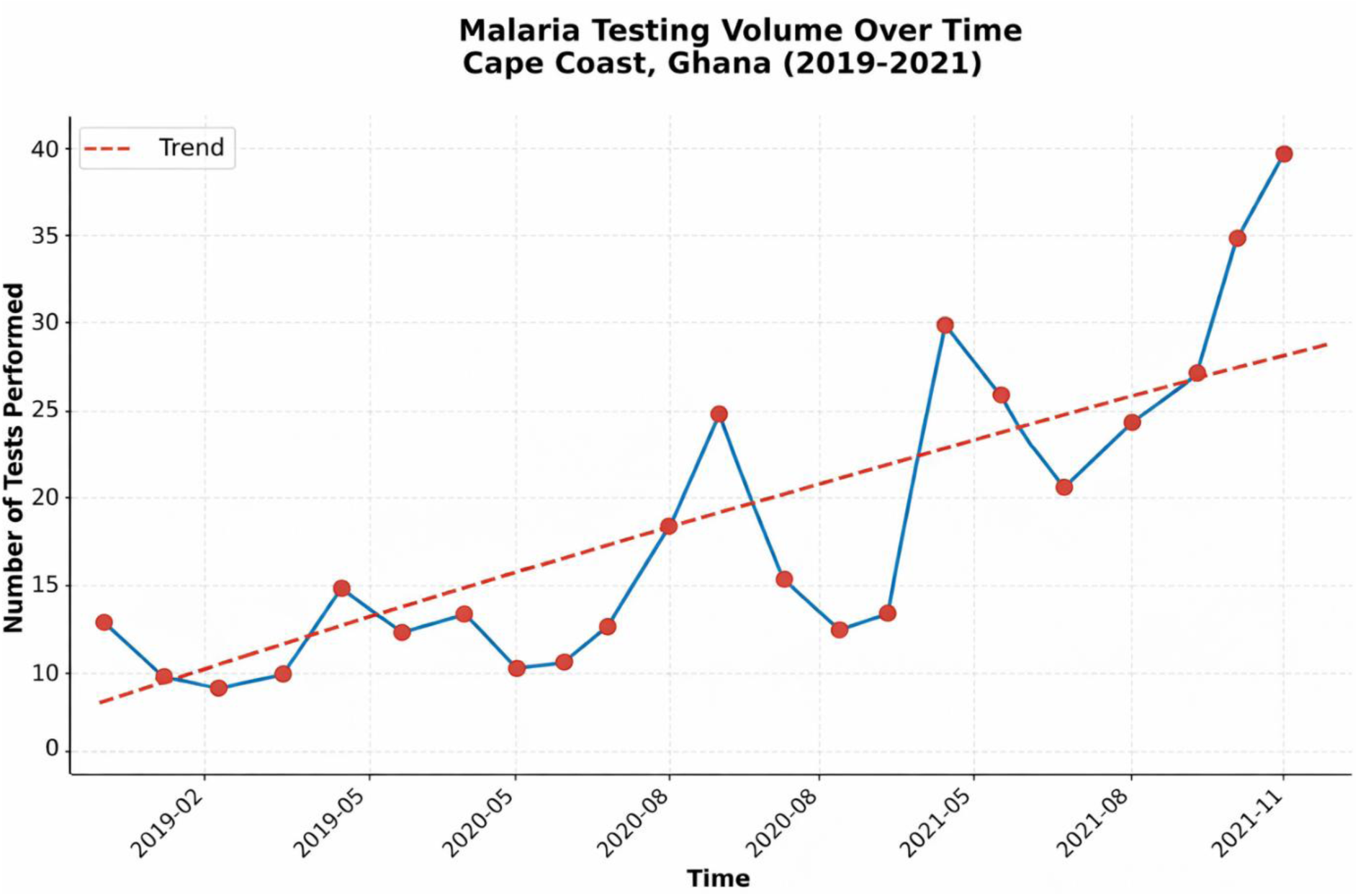
Monthly trend in the number of antenatal visits for hemoglobin testing among pregnant women in Cape Coast, Ghana (January 2019 to December 2021).

### 3.9 Age Distribution

The mean and median age of the pregnant women in the study cohort was 35.0 years (range: 18-45). The distribution was approximately normal, centered on the mean/median value.

**Figure 4:**
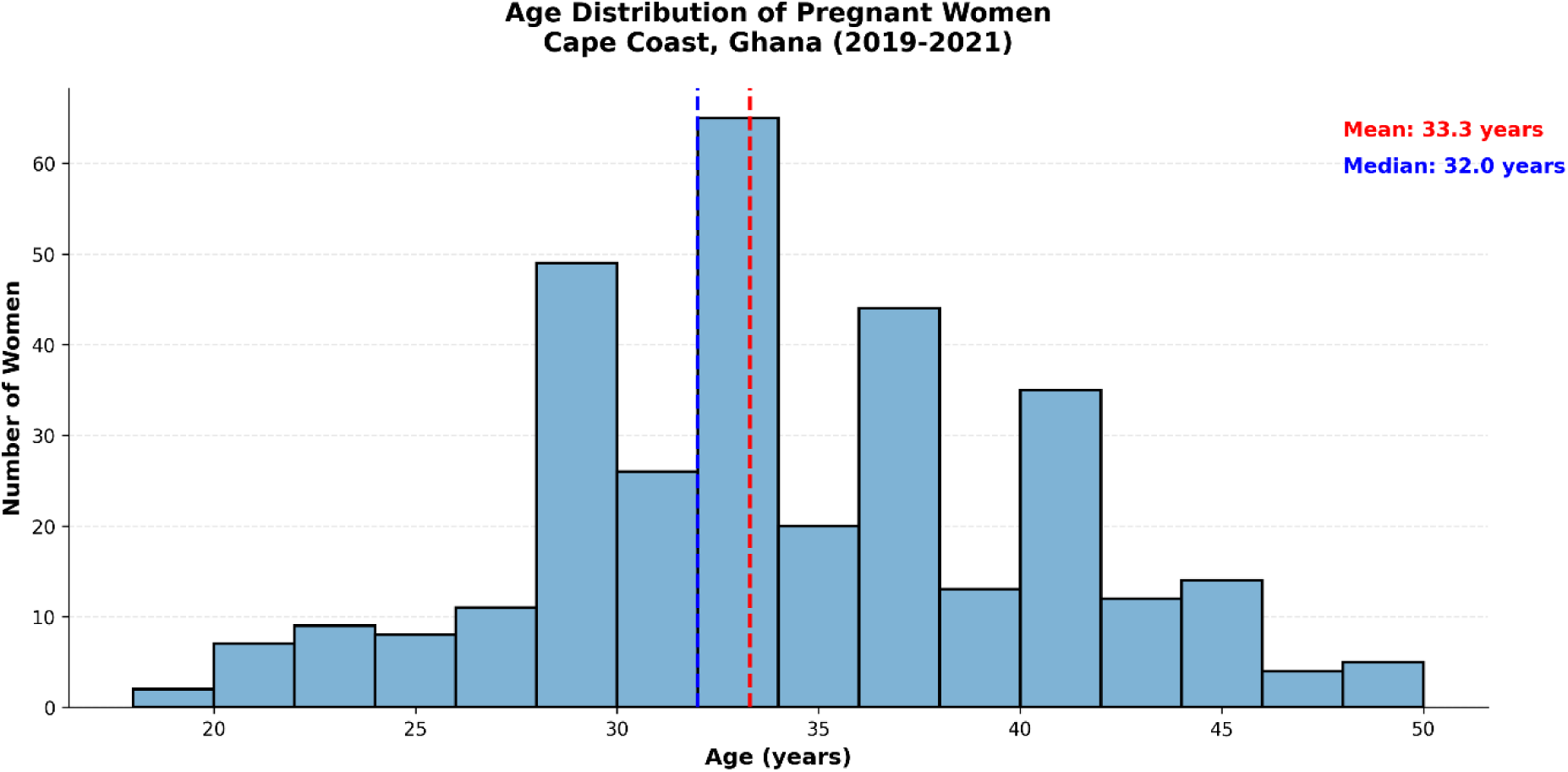
Age distribution of pregnant women in Cape Coast, Ghana (2019–2021), with both the mean and median age of 35.0 years.

### 3.11 Gestational Age at Testing

Gestational age at the time of antenatal hemoglobin testing ranged from 5 to 39 weeks. The distribution indicates testing occurred across all trimesters, with notable frequencies observed at 16 weeks (n=4), 20 weeks (n=3), 28 weeks (n=4), 29 weeks (n=3), and 30 weeks (n=13). The mode of the distribution was 30 weeks.

### 3.12 Quarterly Malaria Prevalence

Malaria prevalence among pregnant women varied by quarter. In 2020, prevalence was highest in Q2 (6.25% in Q2-2020) and Q3 (5.88% in Q3-2020). In 2021, prevalence was 0.0% for the first three quarters (Q1, Q2, Q3) before rising to 4.30% in Q4-2021. An entry marked "non-Qian" also showed a prevalence of 0.0%.

**Figure 5:**
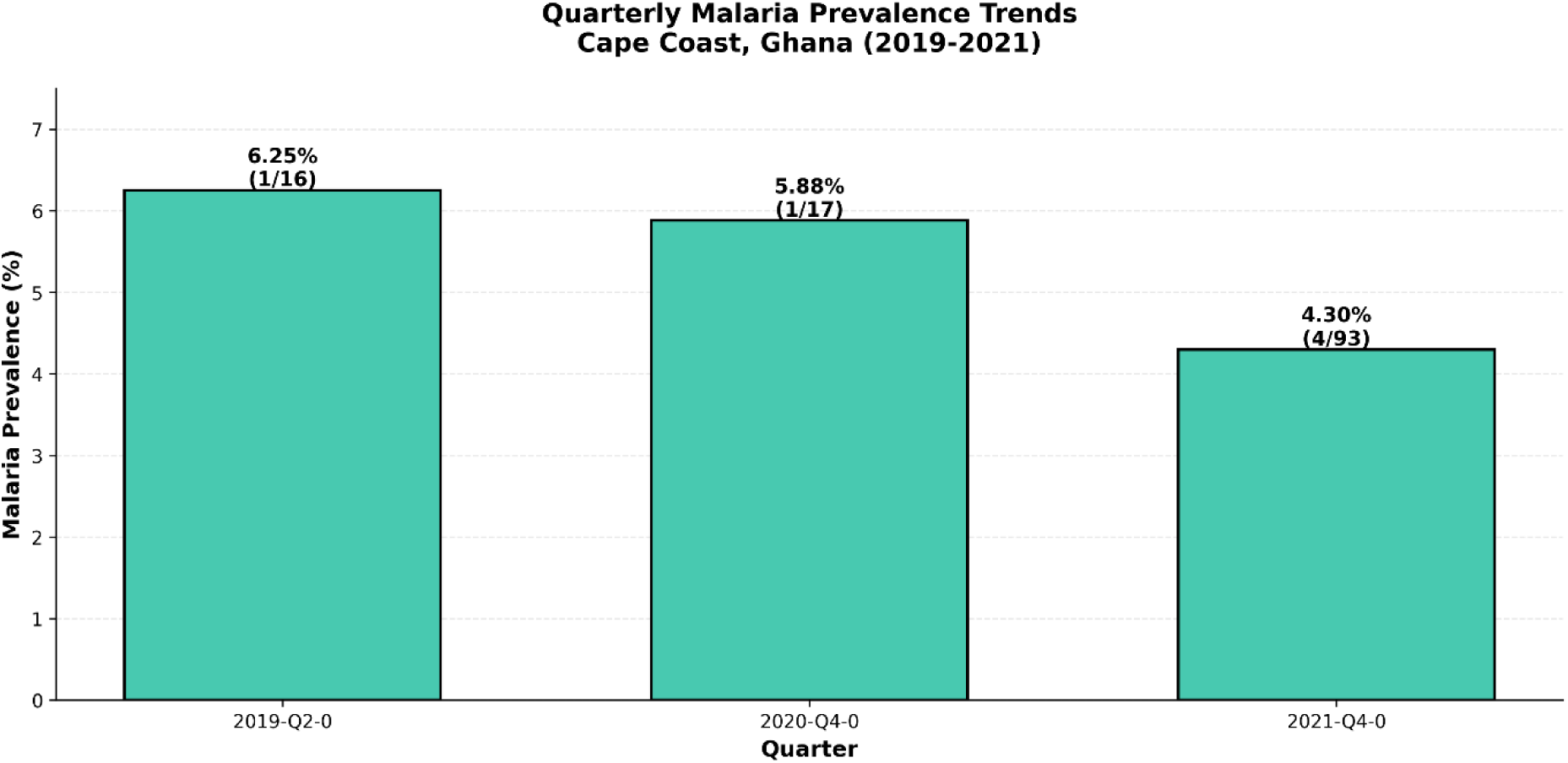
Quarterly malaria prevalence among pregnant women in Cape Coast, Ghana (2019–2021), showing variation in infection rates over time.

### 3.13 Monthly Testing Uptake and Anemia Severity

The monthly number of antenatal hemoglobin tests conducted showed a clear upward trend over the study period (January 2020 to November 2021). Initial counts were low (e.g., n=14 in Jan-2020, n=5 in Feb-2020). After a period of fluctuation, counts increased substantially in 2021, peaking at 39 tests in November 2021. This represents nearly a threefold increase from the first month of observation. Of the 218 women assessed, 101 (46.3%) were anemic (Hb <11 g/dL). Among these, mild anemia (Hb 10-10.9 g/dL) was the most common, affecting 50 women (49.5% of anemic cases). Moderate anemia (Hb 7-9.9 g/dL) was present in 45 women (44.6%), while severe anemia (Hb <7 g/dL) was less frequent, affecting 6 women (5.9%). The majority of the cohort (117 women, 53.7%) had normal hemoglobin levels.

**Figure 6:**
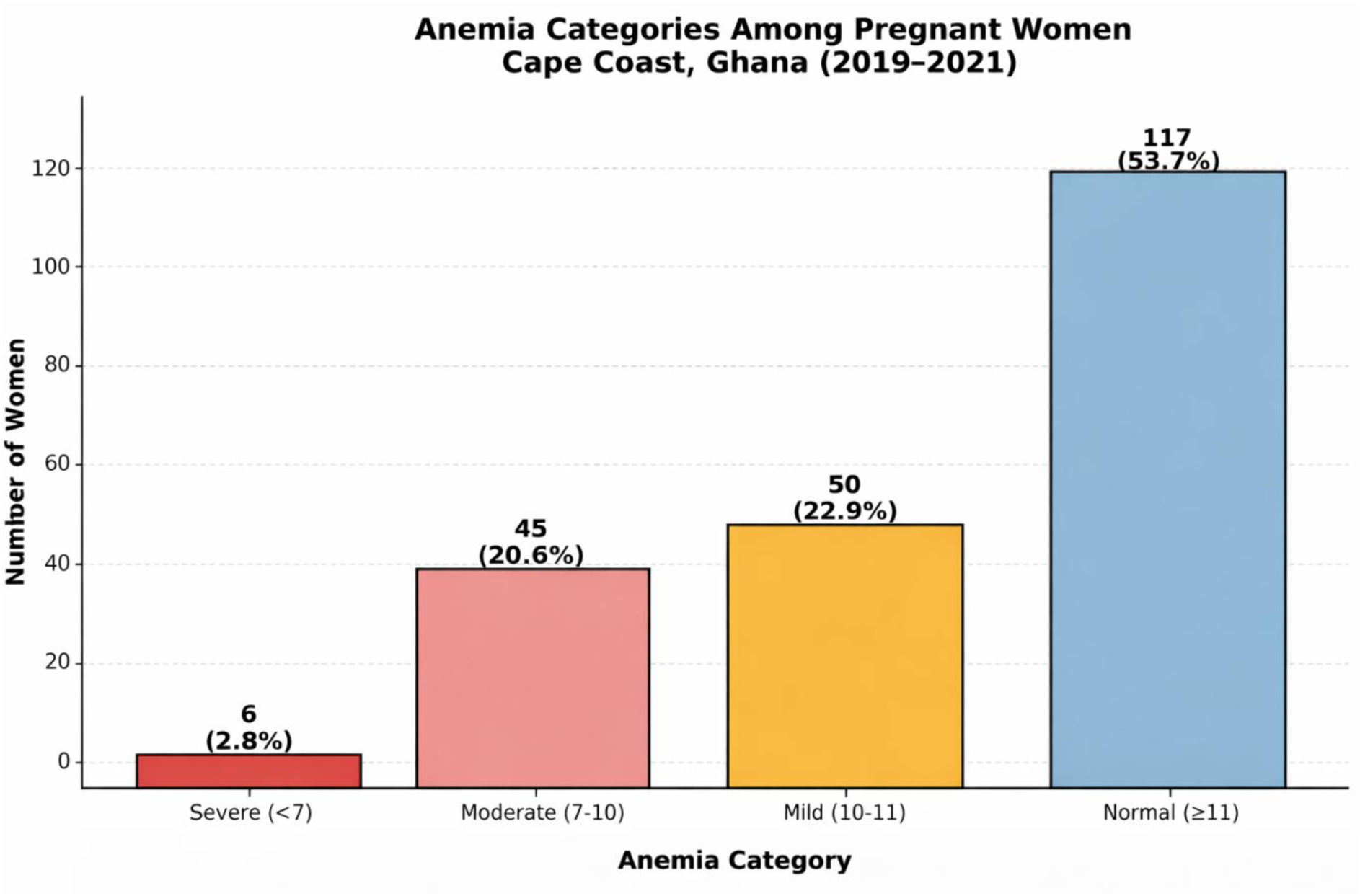
Distribution of anemia severity among pregnant women in Cape Coast, Ghana (2019–2021), with the majority classified as having normal hemoglobin levels.

### 3.14 Association Between Gestational Trimester and Method of Malaria Detection

Among the 191 malaria cases analyzed, the majority were detected using microscopy alone (186 cases), while only 5 cases were identified using a combination of microscopy and rapid diagnostic test (RDT). For cases detected by microscopy, most occurred in the third trimester (136 cases; 73.1%), followed by the first trimester (35 cases; 18.8%) and the second trimester (15 cases; 8.1%). All cases detected using microscopy plus RDT were recorded in the third trimester (5 cases; 100.0%), with no detections in the first or second trimesters using this combined method.

### 3.15 Association Between Hemoglobin Category and Method of Malaria Detection

Among the 191 malaria cases included in the analysis, 186 cases were detected by microscopy alone, while 5 cases were identified using a combination of microscopy and rapid diagnostic test (RDT). Within the microscopy-only group, the majority of cases had normal hemoglobin levels (≥ 11 g/dL) (118 cases; 63.4%). This was followed by moderate anemia (7–9.9 g/dL) (34 cases; 18.3%), mild anemia (10–10.9 g/dL) (29 cases; 15.6%), and severe anemia (< 7 g/dL) (5 cases; 2.7%).

All cases detected using microscopy plus RDT occurred among women with normal hemoglobin levels (5 cases; 100.0%), with no detections in the anemic categories. When totals across detection methods were considered, nearly two-thirds of cases were among women with normal hemoglobin (123 cases; 64.4%), while moderate anemia, mild anemia, and severe anemia accounted for 17.8%, 15.2%, and 2.6%, respectively. Pearson’s chi-square test indicated no statistically significant association between hemoglobin category and method of malaria detection (χ² (3, N = 191) = 2.84, p = 0.417), suggesting that the method of malaria detection did not differ significantly across hemoglobin categories in this sample.

## 4.0 Discussion

### 4.1 Malaria Prevalence Trends

The 2.04% malaria prevalence observed in this study reflects the substantial progress Ghana has made in malaria control over the past two decades. Awine & Silal, 2022) documented a systematic decline in malaria infection among pregnant women across Ghana’s ecological zones, with coastal savannah areas showing reductions from 28-35% in 2003-2010 to 5-11% by 2018-2019. Our findings align with this downward trajectory and suggest continued improvement in the Cape Coast area. Similarly, a recent multicenter study by Bardoe et al. (2024) reported a 10.8% prevalence in the Bono East Region, which, while higher than our findings, still represents improvement from historical rates (Bardoe et al., 2024). The lower prevalence in Cape Coast may reflect the urban-coastal setting, as urban areas have demonstrated more pronounced declines in malaria transmission compared to rural regions.

However, considerable regional variation persists across Ghana. Ampofo et al. (2023) reported malaria prevalence of 17.1% among pregnant women in the Ejisu Municipality of Ashanti Region, substantially higher than our Cape Coast findings. This heterogeneity underscores the importance of subnational surveillance and tailored intervention strategies. Geographic, ecological, and health system factors all contribute to these variations, supporting Ghana’s adoption of a high-burden, high-impact approach targeting areas with persistent transmission.

### 4.2 Seasonal Patterns and Transmission Dynamics

The counterintuitive finding of higher malaria prevalence during the dry season warrants careful interpretation. In Ghana’s coastal and southern regions, rainfall typically follows a bimodal pattern with major rainy seasons from April to June and September to November, and transmission peaks are generally expected 1-2 months after peak rainfall. Our observation of elevated prevalence during November-December (dry season) may reflect delayed transmission from the preceding wet season, as demonstrated by studies showing malaria cases lag rainfall by 1-2 months. Asante et al. (2022) found that in Greater Accra Region, June and July represented peak malaria months following the first major rainy season, while our Q4 clustering suggests residual transmission from the September-November rains.

Alternative explanations for dry season prevalence include microecological factors supporting vector survival during dry periods. Research in other African settings has demonstrated that peri-domestic vegetation and residual water bodies can sustain Anopheles populations during dry seasons. Additionally, testing intensity may have varied seasonally, with increased healthcare-seeking behavior during dry months potentially contributing to detection bias. The clustering of 67% of positive cases in Q4 2021 suggests either a local outbreak event or enhanced surveillance during this period, highlighting the need for continuous year-round malaria screening in pregnancy.

### 4.3 Anemia Burden and Malaria

The 41.3% anemia prevalence observed in our study represents a moderate public health burden according to WHO classifications. This finding is consistent with recent Ghanaian studies: Ampofo et al. (2025) reported 44.5% anemia prevalence in the Ashanti and Volta regions, while Tettegah et al. (2024) found 50% prevalence in the Volta Region. The high anemia burden despite low malaria prevalence indicates that factors beyond malaria - including nutritional deficiencies, hemoglobinopathies, and helminth infections - play predominant roles in maternal anemia in this setting (Ampofo et al., 2022).

The unexpected observation of higher mean hemoglobin in malaria-positive women (13.84 g/dL) compared to negative women (10.90 g/dL) likely reflects the small sample size (n=5 with complete data) and potential selection bias. Ampofo et al. (2025) demonstrated that malaria infection remains an important determinant of maternal anemia even at low prevalence, with infected women showing increased odds of anemia. Our contradictory finding may indicate that asymptomatic parasitemia with preserved hemoglobin status was more readily detected, while severely anemic women may have had unmeasured acute infections treated before testing. Larger prospective studies are needed to clarify the malaria-anemia relationship in low-transmission settings.

### 4.4 Gravidity and malaria risk

Although the study had limited cases for stratified analysis, literature consistently demonstrates that primigravidae and secundigravidae face substantially higher risk of malaria infection during pregnancy compared to multigravidae. This pattern reflects the development of pregnancy-specific immunity to *P. falciparum* antigens, particularly VAR2CSA, which binds to chondroitin sulfate A in the placenta. Studies across Africa have shown that primigravidae have 2-7 times higher risk of placental malaria and adverse outcomes including low birth weight (Rogerson et al., 2018; Umbers et al., 2011).

In our cohort, 34.2% were primigravidae and 65.8% multigravidae, with a mean parity of 2.50 pregnancies. While we could not establish gravidity-specific malaria rates due to small case numbers, this distribution is important for interpreting the low overall prevalence. The higher proportion of multigravidae, who have acquired pregnancy-specific immunity through previous pregnancies, may contribute to the low observed prevalence.

### 4.5 *Plasmodium falciparum* Prevalence

The finding that all positive cases were attributable to *P. falciparum* is consistent with Ghana’s malaria epidemiology, where *P. falciparum* accounts for over 90% of infections (Mwin et al., 2021). This species poses particular risks during pregnancy through its ability to sequester in the placental microcirculation, causing placental insufficiency, maternal anemia, and fetal growth restriction (Obiri et al., 2020). The exclusive detection of *P. falciparum* underscores the continued relevance of pregnancy-specific interventions targeting this parasite.

### 4.6 Implications for Public Health Practice

#### Malaria Prevention Strategies

The low malaria prevalence documented in this study supports Ghana’s ongoing transition toward elimination, consistent with the *National Malaria Elimination Strategic Plan 2024–2028* (Ghana Health Service/NMCP, 2024). The plan outlines Ghana’s goal of achieving malaria elimination by 2030 through intensified surveillance, universal coverage of insecticide-treated nets (ITNs), and high uptake of intermittent preventive treatment in pregnancy with sulfadoxine-pyrimethamine (IPTp-SP). Evidence from recent studies confirms the effectiveness of these strategies. Ampofo et al. (2025) demonstrated that adherence to IPTp-SP and consistent ITN use substantially reduced malaria prevalence and anemia risk among pregnant women in Weija-Gbawe District (medRxiv preprint). Similarly, Ansong et al. (2025) observed that IPTp-SP uptake in Ashanti Region remains a cornerstone of malaria control, showing measurable protection against infection and maternal anemia. The national ITN distribution program has also proven resilient, maintaining high coverage during and after the COVID-19 pandemic (Nuñez et al., 2023).

Despite these gains, the persistent detection of *Plasmodium falciparum* infections highlights the need for sustained vigilance. The unexpected dry-season prevalence observed in our study suggests that malaria screening and prevention should not be restricted to traditional rainy-season peaks. Continuous, year-round screening of pregnant women during all antenatal contacts which are aligned with the WHO’s eight-contact model for early detection and treatment. In low-transmission settings such as urban coastal Ghana, complacency could reverse hard-won progress; therefore, ongoing health worker training, real-time surveillance, and routine IPTp-SP monitoring remain essential.

#### Anemia Control in Low-Transmission Settings

The disproportionately high anemia burden relative to malaria prevalence in this study underscores the multifactorial nature of maternal anemia. Recent analyses from the 2019 Ghana Malaria Indicator Survey identified nutritional iron and folate deficiencies, intestinal parasitic infections, and socioeconomic inequities as the dominant predictors of anemia among pregnant women (Klu et al., 2024). While malaria prevention remains an important pillar, interventions should concurrently target nutritional and social determinants. Ghana’s focused antenatal care model which assigns pregnant women to a consistent provider offers an excellent framework for integrating dietary counseling, iron-folic acid supplementation, and routine hemoglobin monitoring. Aballo et al. (2025) reported that trimester-specific anemia screening and supplementation significantly improved birth outcomes in Ghanaian health facilities (Aballo et al., 2025). The 41.3 % anemia prevalence in this study indicates that existing measures, though beneficial, require intensification. Strengthened nutrition counseling emphasizing iron-rich foods, improved supplementation adherence, and early detection of severe anemia (< 7 g/dL) should be prioritized to prevent maternal and perinatal complications.

### 4.7 Limitations

The small number of malaria-positive cases (n=6) substantially limits statistical power for comparative analyses and precludes meaningful multivariable modeling of risk factors.

Again, substantial missing data for key variables including gestational age (39.8% missing), hemoglobin (34.4% missing), and educational level (99.1% missing) reduces analytical capacity and may introduce selection bias.

Furthermore, the reliance on facility-based sampling may overestimate preventive intervention coverage and underestimate malaria burden if women with barriers to care are underrepresented.

Finally, the absence of molecular diagnostic methods means submicroscopic parasitemia, which contributes to ongoing transmission, was not assessed.

The study’s restriction to a single urban facility in the Central Region limits generalizability to rural settings and other ecological zones. Malaria epidemiology varies substantially across Ghana’s three major zones - northern savannah, middle transitional/forest, and coastal savannah with different transmission patterns and intervention coverage. Future multicenter studies incorporating both urban and rural sites across multiple regions would provide more comprehensive national perspectives on malaria in pregnancy burden and trends.

The small number of malaria-positive cases (n=6) substantially limited our analytical capacity, precluding meaningful stratified analyses by risk factors such as gravidity, trimester, or anemia status. While sensitivity analyses demonstrated that the overall prevalence estimate was reasonably robust (ranging from 2.04% to 6.12% across different sample definitions with overlapping confidence intervals), the wide confidence intervals reflect the limited precision inherent in small case numbers.

The substantial missing data for gestational age (61.7%) and hemoglobin (34.7%) reduced the effective sample size for complete case analysis to 98 participants (29.3% of total). However, missing data appeared to occur at random rather than systematically, minimizing potential for selection bias. The paradoxical observation of higher hemoglobin in malaria-positive women should be interpreted with extreme caution given the small sample size (n=6) and likely reflects selection bias or chance variation rather than a true biological relationship.

Future multicenter studies with larger sample sizes, more complete data collection, and prospective design would enable more robust analyses of risk factors, pregnancy outcomes, and the malaria-anemia relationship in low-transmission settings approaching elimination.4

### 4.9 Recommendations for Future Research

Several critical research gaps emerged from this study.

First, larger multicenter studies with adequate sample sizes are needed to robustly characterize seasonal patterns and identify true risk factors for malaria in low-transmission settings. Pooling data from multiple coastal facilities could provide sufficient power for meaningful analyses while maintaining ecological comparability.

Second, incorporation of molecular diagnostic methods (PCR) would enable detection of submicroscopic parasitemia, which persists even as microscopic prevalence declines and contributes to ongoing transmission.

Third, pregnancy outcome data including birth weight, gestational age at delivery, and adverse events should be systematically collected to assess the clinical significance of low-level malaria parasitemia and anemia in contemporary Ghana. Understanding whether the health impacts of malaria have changed as transmission has declined would inform prevention strategy priorities. Fourth, comprehensive assessment of IPTp-SP uptake, insecticide-treated net usage, and adherence to antenatal care recommendations would help identify gaps in preventive intervention coverage and guide quality improvement efforts.

Finally, qualitative research exploring healthcare-seeking behaviors, community perceptions of malaria risk in low-transmission settings, and barriers to antenatal care attendance would provide crucial context for designing acceptable and effective interventions.

## 5.0 Conclusions

This study documents continued progress in malaria control among pregnant women in Cape Coast, Ghana, with prevalence declining to 2.04% during 2019-2021. This finding supports Ghana’s trajectory toward malaria elimination while highlighting that transmission has not ceased and vigilance must be maintained. The unexpected seasonal pattern with higher dry season prevalence suggests that year-round screening and prevention are essential, and transmission dynamics in low-burden settings suggest further investigation.

The persistent high anemia burden despite low malaria prevalence indicates that non-malarial causes of maternal anemia require intensified attention. Comprehensive antenatal care addressing nutritional deficiencies, early hemoglobin screening, and targeted supplementation should be prioritized alongside continued malaria prevention. As Ghana progresses toward malaria elimination, health systems must adapt strategies to maintain prevention in low-transmission contexts while addressing the broader spectrum of maternal health challenges.

Continued surveillance, sustained implementation of evidence-based preventive interventions, and research to optimize strategies for low-transmission settings will be essential to achieving and maintaining malaria elimination goals while improving overall maternal and child health outcomes in Ghana.

## AI STATEMENT

ChatGPT version 5.2 (OpenAI) and other artificial intelligence techniques were only utilized for document organization and language refining. Data collection, data analysis, result interpretation, and figure development were all done without the help of AI. All AI-assisted content was reviewed, revised, and validated by the authors, who also accept full responsibility for the manuscript’s correctness and integrity.

## Data Availability

All data produced in the present study are available upon reasonable request to the authors

## Notes

### Competing Interest Statement

The authors have declared no competing interest.

### Funding Statement

This study did not receive any funding

### Author Declarations

The study used (or will use) ONLY openly available human data that were originally located at the Cape Coast Teaching Hospital. Ethical approval was obtained from the Cape Coast Teaching Hospital Ethical Review Committee (CCTHERC/EC/2025/182)

